# Bayesian Estimation Improves Prediction of Outcomes after Epilepsy Surgery

**DOI:** 10.1101/2024.06.21.24309313

**Authors:** Adam S. Dickey, Vineet Reddy, Nigel P. Pedersen, Robert T. Krafty

## Abstract

Low power is a problem in many fields, as underpowered studies that find a statistically significant result will exaggerate the magnitude of the observed effect size. We quantified the statistical power and magnitude error of studies of epilepsy surgery outcomes. The median power across all studies was 14%. Studies with a median sample size or less (n<=56) and a statistically significant result exaggerated the true effect size by a factor of 5.4 (median odds ratio 9.3 vs. median true odds ratio 1.7), while the Bayesian estimate of the odds ratio only exaggerated the true effect size by a factor of 1.6 (2.7 vs. 1.7). We conclude that Bayesian estimation of odds ratio attenuates the exaggeration of significant effect sizes in underpowered studies. This approach could help improve patient counseling about the chance of seizure freedom after epilepsy surgery.

**SHORT SUMMARY:** We estimated the statistical power of studies predicting seizure freedom after epilepsy surgery. We exacted data from a Cochrane meta-analysis. The median power across all studies was 14%. Studies with a median sample size or less (n<=56) and a statistically significant result exaggerated the true effect size by a factor of 5.4, while the Bayesian estimate of the odds ratio only exaggerated the true effect size by a factor of 1.6. We conclude that Bayesian estimation of odds ratios attenuates the exaggeration of significant effect sizes in underpowered studies. This result could improve patient counseling regarding epilepsy surgery.

## INTRODUCTION

Low statistical power is a recognized problem in many fields^1^, which means true effects will be missed, the false discovery rate will be inflated^2^, and discovered true effects will be over-estimated^3, 4^. We highlighted the issue of low power in a single study of minimally invasive surgery^5^, but suspected that low power was pervasive in the field of epilepsy surgery. In this study, we estimate the median statistical power of studies of epilepsy surgery outcomes. We argue that Bayesian estimation can attenuate the exaggeration of statistically significant effect sizes.

In neurosciences, the median power to detect a true effect was estimated at 20%^6^. That study defined “ground truth” as the pooled effect size of a meta-analysis, and then calculated the statistical power of each included study to find that pooled effect. We apply that same approach to the Cochrane Library Review of Surgery for Epilepsy, which included data from 182 studies with 16,855 participants^7^.

A Bayesian approach can prevent overfitting in small sample sizes by constraining estimates with prior knowledge. A prior distribution is combined with the likelihood of the observed data to create an optimal posterior estimate. In this study, we use Bayesian logistic regression. The slope of a logistic regression can be interpreted as the log of the odds ratio for a prognostic factor. The Bayesian approach requires that prior knowledge be formalized as distributions of plausible slopes and intercepts. These prior distributions should be independent of the observed data.

We quantified our prior knowledge using an separate study from the European Epilepsy Brain Bank, which reported 1-year seizure free outcomes for 8,247 participants organized by histopathological diagnosis^8^. We performed pairwise comparisons of these diagnoses to quantify a prior distribution of plausible odds ratios. We then used this prior distribution in Bayesian logistic regression on data from the Cochrane review. Finally, we computed the error between the “ground truth” odds ratio and the observed odds ratio, compared to the error between ground truth and the Bayesian estimate of the odds ratio.

## METHODS

### Cochrane Data

We extracted publication level data from “Comparison 4. Prognostic factors of a good outcome of surgery” from the Cochrane review^7^, including the number of participants and number with a good outcome (seizure free or Engel I) for both the experimental and control group. We recalculated the meta-analysis to create a pooled odds ratio, rather than the reported risk ratio, using the package *meta* in statistical software R (version 4.2.1). The fixed effect estimate for each comparison was used as ground-truth for all included studies. We excluded three analyses which duplicated studies from a previous analysis (comparisons of 4.13, 4.15, and 4.17).

### Power analysis

Our “ground truth” effect size was derived for each prognostic factor by taking the percentage of participants with a good outcome in the control group, and applying the pooled odds ratio to derive the percentage with good outcome in the experimental group. We computed the statistical power (using R package *exact*) to detect that ground truth effect size for each study for a given prognostic factor, assuming a Chi-squared test. We reported the median power for all studies whose pooled effect was statistically significant in the original Cochrane study.

### Bayesian estimation and prior distribution

The prior distribution for plausible slopes was derived from the reported 1-year seizure freedom by histopathological diagnosis in the European Epilepsy Brain bank study^8^. We performed all possible pairwise comparisons of 17 non-overlapping categories that had at least 100 participants. We assumed zero mean and computed the standard deviation of the log-odds ratios of the pairwise comparisons to define a Gaussian prior distribution for slope. The prior for the intercept was weakly informative (Gaussian with zero mean, standard deviation 1000). We used these prior distributions to perform Bayesian logistic regression using the probabilistic language Stan^9^ (R package *rstanarm*). We computed the mean squared error of the observed log odds ratio from the ‘ground truth” log odds ratio and compared this to the error of the Bayesian log odds ratio from the “ground truth” log odds ratio using a paired t-test.

### Exaggeration of statistically significant results

We examined the relationship of the sample size to the magnitude of the observed odds ratio in studies which were statistically significant (Chi-squared test with p-value < 0.05) and in the same direction as the “ground truth” odds ratio. We added 0.5 to all cells if there was a zero cell value. We compared this to the relationship of the sample size to the magnitude of the Bayesian odds ratio in studies where the posterior probability of an odds ratio > 0 was more than 95%, and in the same direction as “ground truth.” We estimated the exaggeration ratio between the observed versus the Bayesian odds ratio and the median true odds ratio for studies with sample size less than or equal to the median.

## RESULTS

We extracted data from 367 binary comparisons of good outcome after epilepsy surgery according to one of 14 prognostic factors. We flipped all odds ratios to be positive, so the experimental group was associated with better chance of good outcome. The “ground truth” odds ratios ranged from 1.05 (positive history of head injury vs. no such history) to 2.54 (complete resection vs. less complete). The median “ground truth” odds ratio across all studies was 1.72 (for presence of mesial temporal sclerosis vs. not).

The median sample size was 56, with an inter-quartile range (IQR) of 42 to 100. We computed the statistical power of each comparison to detect the pooled effect side for the prognostic factors whose meta-analysis showed a significant result. The median power across all studies was 14% (IQR 10% to 23%). Low power is known to increase the chance a study is a false positive^10^. If the pre-test probability of study being true is 50%, then the chance is study with 14% power and a significant result (at p < 0.05) is a false positive is 0.05 / (0.05+0.14) = 26%.

We derived a prior distribution of odds ratios from the European Brain bank study. The prior distribution for plausible slopes (i.e. log odds ratios) was a normal distribution with mean zero and standard deviation (STD) of 0.70 (Figure 1A). This implies that 68% of odds ratios are between ½ and 2 and 95% are between ¼ and 4. We found that this qualitatively fit the observed log odds ratios for large (sample size >= 100) studies from the Cochrane review, with two exceptions. First, one outlying study reported 69/79 patients seizure free with complete resection and 3/28 for less complete resection, for observed odds ratio of 57.5^11^. However, outliers are expected, given that 107 participants in still underpowered to detect a small effect size. Second, there are few studies with log odds ratio around 0. This is consistent with publication bias, as researchers are less likely to write up null findings^12^.

**Figure 1:**
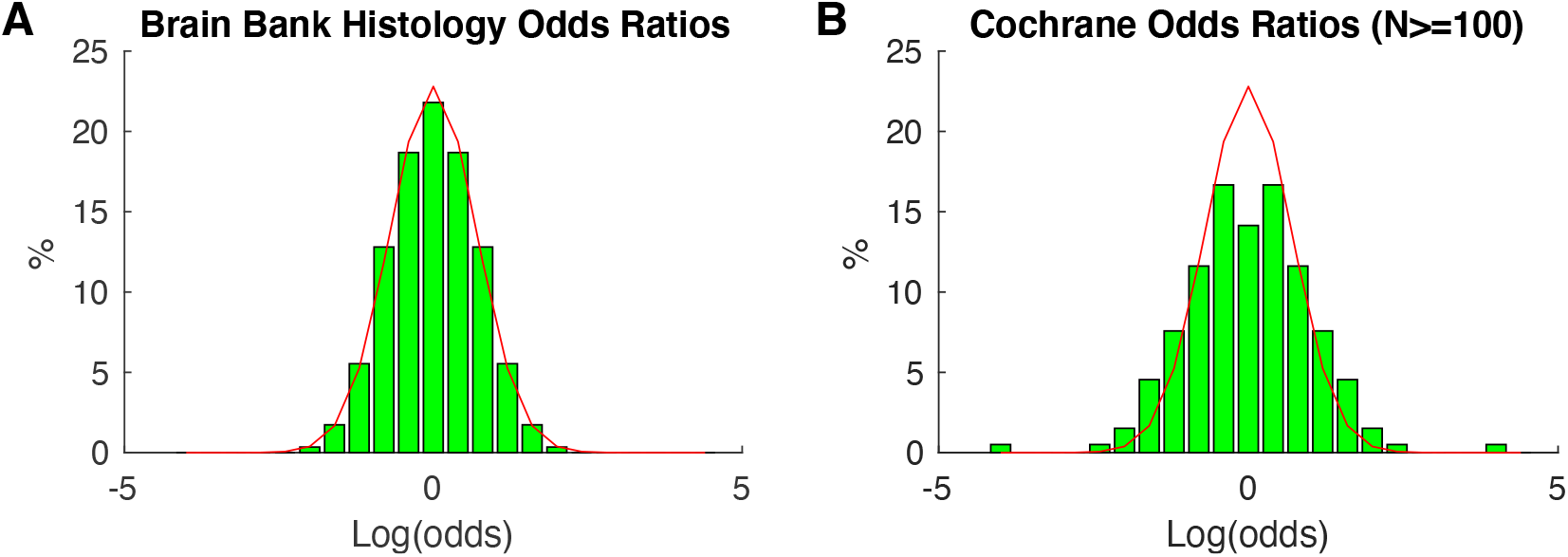
Prior distribution of log odds ratios. **A)** The prior distribution for the log odds ratio, derived from the European Epilepsy Brain Bank, was normally distribution with mean zero and standard deviation of 0.7 (red). This implies that 95% of the odds ratios are between ¼ and 4. **B)** This prior distribution (red) qualitatively matches the observed log odds ratios from large studies (sample size >= 100) from the Cochrane review. All odds ratios are plotted twice to include the reciprocal of the observed odds ratios. There is one outlier and a paucity of null studies with log odds ratio of 0 (i.e odds ratio of 1).

We assumed a weakly-informative prior distribution for the intercept (see Methods). Given prior distributions for slope and intercept, we can derive the Bayesian estimate of the log odds ratio using a probabilistic statistics package^9^. The mean squared error of the observed log odds ratio from the “ground truth” odds ratio (1.0) was significantly higher (p<0.001, paired t-test) than the mean squared error of the Bayesian log odds ratio to “ground truth” (0.27).

An underpowered study with a small sample size and a statically significant effect will over-estimate the true effect size. We compared the observed and Bayesian odds ratios as a function of sample size (Figure 2). We found that a study with a median sample size or less (n<=56) and a statistically significant result exaggerated the true effect size by a factor of 5.4 (median odds ratio 9.3 vs. median true odds ratio 1.7). In contrast, a Bayesian estimate of odds ratio with a neutral prior only exaggerated the true effect size by factor of 1.6 (2.7 vs. 1.7).

**Figure 2:**
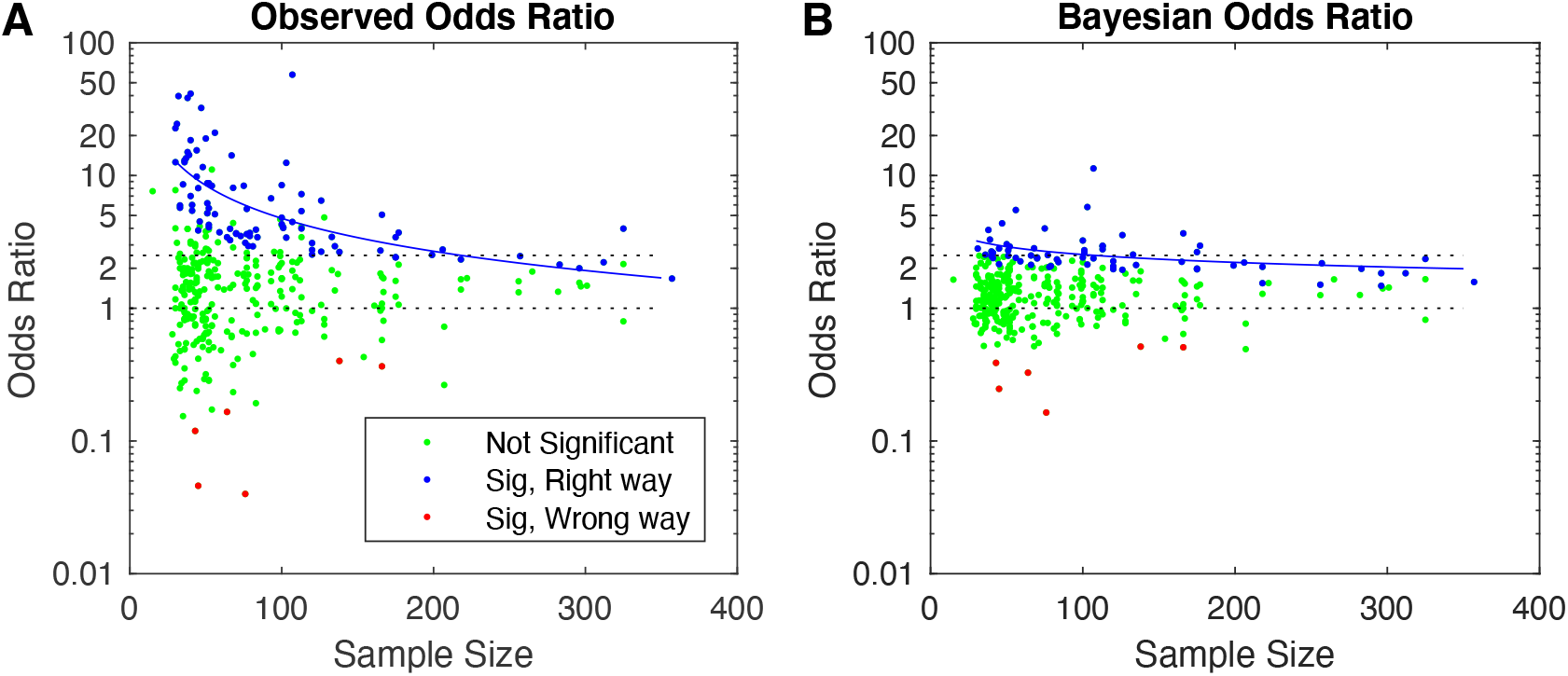
Observed versus Bayesian odds ratios. **A)** Observed odds ratios are shown as a function of sample size on a semi-log plot. The median true odds ratio was 1.7, and ranged from 1 to 2.5 (black dotted lines). Observed odds ratios from studies that are statistically significant (Chi-squared p-value < 0.05) and in the same direction of the true effect (blue dots) over-estimate the true odds ratio at low sample sizes, as shown by a quadratic trend line (blue line) **B)** Bayesian odds ratios are shown as a function of sample size on a semi-log plot. The Bayesian estimate of odds ratios from significant studies (greater than 95% posterior probability of odds ratio above 1) that are also in the same direction of the true odds ratio (blue dots) show less over-estimation.

## DISCUSSION

In this study, we found that the median power of a typical study of epilepsy surgery was only 14%. We showed that studies with small sample sizes and statistically significant results exaggerated the magnitude of observed effect. We showed that Bayesian estimation of the odds ratio can attenuate this exaggeration of significant effect sizes in underpowered studies.

We believe Bayesian estimation is relevant for clinicians counseling patients about epilepsy surgery. A recent paper examined whether low voltage fast activity (LVFA) intracranial ictal onset predicted seizure freedom after minimally invasive surgery: they reported 46% (20/43) chance of seizure freedom with LVFA, but 0% (0/15) chance without LVFA^13^. That observed odds ratio is infinite. A corrected odds ratio (adding 0.5 to each cell) is (20.5 / 23.5) / (0.5 / 15.5) = 27, which presumably exaggerates the true effect size. The Bayesian estimate of the odds ratio is 3.6. The value is much closer to the observed odds ratio of 3.3 from a larger study of LVFA– 61% (68/114) seizure free with vs. 31% (9/29) without^14^. This suggests that lack of a LVFA onset lowers the chance of seizure freedom, but not to zero. Surgery might still be a reasonable option, based on the remaining clinical data and the patient’s preferences.

The main limitation of this approach is that Bayesian logistic regression does not have an analytical solution, which makes it less accessible. State of the art statistical packages are probabilistic, utilizing Markov Chain Monte Carlo methods^9^. Another limitation is that our prior distributions are narrowly defined for predicting seizure freedom after epilepsy surgery. Prediction of another outcome (such as risk of cognitive decline) would require a separate derivation of prior distributions.

There are alternatives for shrinking observed odds ratios to a plausible range, including ridge regression, LASSO, and elastic net^15^. While these methods are commonly used, the choice and degree of shrinkage is itself a hyper-parameter that must be optimized. The advantage of the Bayesian approach is that the hyper-parameters (the prior distributions) were derived directly from existing literature. Whatever the method used, we believe it is critical to shrink observed effect sizes from small studies of surgery outcomes. The Bayesian method highlighted here provides clinician a nuanced approach for interpreting results from small studies, with the goal of improving the counseling we give our patients about epilepsy surgery.

## Data Availability

The extracted raw data and an RMarkdown file code which can be used to reproduce the analyses and figures described here will be posted upon publication in a peer reviewed journal at https://github.com/AdamSDickey

## FUNDING AND ACKNOWLEDGEMENTS

N.P.P. was supported by the Woodruff Foundation, CURE Epilepsy, and NIH grants K08 NS105929, R01 NS088748, and R21 NS122011. A.S.D. was supported by the National Center for Advancing Translational Sciences of the NIH under award number UL1 TR002378 and KL2 TR002381. R.T.K. was supported by R01 GM113243. The content is solely the responsibility of the authors and does not necessarily represent the official views of the National Institutes of Health

## DISCLOSURE OF CONFLICTS OF INTEREST

N.P.P. has served as a paid consultant for DIXI Medical USA, who manufactures products used in the workup for epilepsy surgery. The terms of this arrangement have been reviewed and approved by Emory University in accordance with its conflict-of-interest policies. A.S.D, V.R. and R.T.K. have no conflicts of interest to disclose.

## DATA AVAILABILITY

The study used only openly available human data that were previously published by the Cochrane Library^7^ and the European Epilepsy Brain Bank^8^. The extracted raw data and an RMarkdown file code which can be used to reproduce the analyses and figures described here will be posted upon publication in a peer reviewed journal at https://github.com/AdamSDickey

## Notes

### Author Declarations

The study used only openly available human data that were previously published by the Cochrane Library (West et al., 2019) and the European Epilepsy Brain Bank (Lamberink et al., 2020).

## REFERENCES

1. Cohen J. The statistical power of abnormal-social psychological research: A review The Journal of Abnormal and Social Psychology. 1962 1962/09//;65:145–153.

2. Ioannidis JPA. Why Most Published Research Findings Are False PLoS Med. 2005 2005/08/30/;2:e124.

3. Ioannidis JPA. Why Most Discovered True Associations Are Inflated Epidemiology. 2008 2008/09//;19:640–648.

4. Gelman A, Carlin J. Beyond Power Calculations: Assessing Type S (Sign) and Type M (Magnitude) Errors Perspect Psychol Sci. 2014 2014/11//;9:641–651.

5. Dickey AS, Pedersen NP. Low statistical power in a study predicting seizure outcome Epilepsia. 2021 Oct;62:2565–2566.

6. Button KS, Ioannidis JP, Mokrysz C, Nosek BA, Flint J, Robinson ES, et al. Power failure: why small sample size undermines the reliability of neuroscience Nat Rev Neurosci. 2013 May;14:365–376.

7. West S, Nevitt SJ, Cotton J, Gandhi S, Weston J, Sudan A, et al. Surgery for epilepsy Cochrane Database of Systematic Reviews. 2019 2019/06/25/.

8. Lamberink HJ, Otte WM, Blümcke I, Braun KPJ, Aichholzer M, Amorim I, et al. Seizure outcome and use of antiepileptic drugs after epilepsy surgery according to histopathological diagnosis: a retrospective multicentre cohort study The Lancet Neurology. 2020 2020/09//;19:748–757.

9. Carpenter B, Gelman A, Hoffman MD, Lee D, Goodrich B, Betancourt M, et al. <i>Stan</i> : A Probabilistic Programming Language J Stat Soft. 2017 2017;76.

10. Button KS, Ioannidis JPA, Mokrysz C, Nosek BA, Flint J, Robinson ESJ, et al. Power failure: why small sample size undermines the reliability of neuroscience Nat Rev Neurosci. 2013 2013/05//;14:365–376.

11. Terra-Bustamante VC, Fernandes RMF, Inuzuka LM, Velasco TR, Alexandre V, Wichert-Ana L, et al. Surgically amenable epilepsies in children and adolescents: clinical, imaging, electrophysiological, and post-surgical outcome data Childs Nerv Syst. 2005 2005/07//;21:546–551.

12. Franco A, Malhotra N, Simonovits G. Publication bias in the social sciences: Unlocking the file drawer Science. 2014 2014/09/19/;345:1502–1505.

13. Michalak AJ, Greenblatt A, Wu S, Tobochnik S, Dave H, Raghupathi R, et al. Seizure onset patterns predict outcome after stereo-electroencephalography-guided laser amygdalohippocampotomy Epilepsia. 2023 2023/04/24/:epi.17602.

14. Lagarde S, Buzori S, Trebuchon A, Carron R, Scavarda D, Milh M, et al. The repertoire of seizure onset patterns in human focal epilepsies: Determinants and prognostic values Epilepsia. 2019 2019/01//;60:85–95.

15. Zou H, Hastie T. Regularization and Variable Selection Via the Elastic Net Journal of the Royal Statistical Society Series B: Statistical Methodology. 2005 2005/04/01/;67:301–320.

